# Comparative Effectiveness of Standard and Contingency-based Cleaning in Acute and Long-Term Care Facilities amidst Staff Shortages and a COVID-19 Surge

**DOI:** 10.1101/2021.04.13.21255427

**Authors:** Emil Lesho, Donna Newhart, Lisa Reno, Scott Sleeper, Julia Nary, Jennifer Gutowski, Stephanie Yu, Edward Walsh, Roberto Vargas, Dawn Riedy, Robert Mayo

**Author notes:** Corresponding Author: Emil Lesho, 1425 Portland Avenue, Rochester, NY 14621.

## Abstract

**Background:** Cleanliness of hospital surfaces helps prevent healthcare-associated infections, but larger evaluations of the effectiveness of various cleaning strategies during SARS-CoV-2 surges and worker shortages are scarce.

**Methods:** In an acute care hospital (ACH) and a long-term care facility (LTCF), 417 surfaces were tested for SARS-CoV-2 and adenosine triphosphate before and after various cleaning strategies, including ultraviolet light (UV-C), electrostatic spraying, and room fogging.

**Results:** ACH surface contamination differed among outbreak and non-outbreak wards (p = 0.001). RNA was detected on 66% of surfaces before cleaning and on 23% of those surfaces immediately after terminal cleaning, for a 65% post-cleaning reduction (p = 0.001). UV-C resulted in an 87% reduction (p = 0.023), while spraying with electrostatic bleach resulted in a 47% reduction (p = 0.010). LTCF contamination rates differed between the dementia, rehabilitation, and the residential units (p = 0.005). 67% of surfaces had RNA after room fogging without terminal-style wiping. Fogging with wiping led to an 11% reduction in the proportion of positive surfaces.

**Discussion:** Baseline contamination varied by type of unit and outbreak conditions, but not facility type. Removal of viral RNA varied according to strategy. Unlike previous reports, time spent cleaning was associated with cleaning thoroughness.

## Introduction

Cleanliness of hospital surfaces helps prevent healthcare-associated infections [1, 2]. Contamination of environmental surfaces with SARS-CoV-2 has been reported from various countries, under experimental conditions in the laboratory, or in facilities with bio-containment units [3-11]. However, larger, controlled, pre and post cleaning evaluations, particularly from U.S. health care facilities dealing with nosocomial outbreaks during the height of the pandemic are scarce.

Healthcare associated (HA) transmission of SARS-CoV-2 results from of a complex interplay of several factors, including patient census, nurse-to-patient ratio, adherence to isolation guidelines and policies for using personal protective equipment, patient acuity, and the prevalence of presymptomatic/asymptomatic carriers. However, we hypothesize that these factors may not fully account for large or prolonged nosocomial outbreaks. Environmental contamination could also be an overlooked contributor to HA-transmission.

In addition, the shortage of environmental service (EVS) workers and their concerns regarding cleaning rooms of patients infected with SARS-CoV-2 further complicated cleaning and disinfection efforts [12-15]. This forced some healthcare facilities to use contingency-based approaches such as professional remediation companies. Moreover, long-term care facilities (LTCF) have experienced inordinately high infection and mortality rates [16-18]. The potential contribution of environmental contamination with SARS-CoV-2 to this is uncertain.

Adding to the challenge of investigating the role of environmental SARS-CoV-2 is that fact that the vast majority of studies, including this one, do not have the biosafety level necessary for the cultivation of live virus and must rely on viral RNA as a surrogate. Even though nucleic acid isolated from healthcare surfaces may not represent infectious or cultivable virus, and the infectious potential of environmental nucleic acid, including RNA, is not fully understood, we have shown that environmental nucleic acid from another common pathogen was correlated with nosocomial infections of that species [19].

Our objectives were to assess baseline environmental contamination with SAR-CoV-2 RNA at an acute-care hospital (ACH) and a LTCF and compare contamination rates between different wards or units and between outbreak and non-outbreak settings. Attempting to maintain cleaning standards and quality measures in the midst of critical environmental service worker shortages, the LTCF hired a commercial cleaning and remediation company as a contingency measure. Therefore, we also sought to evaluate the effectiveness of that approach.

## Settings

The ACH is a 528-bed teaching hospital in the Finger Lakes Region of New York State. The baseline configuration pre-pandemic surge is such that 42% of the rooms are semi-private. There are 48 intensive care beds and 14 special care nursery beds. It has the eleventh busiest emergency department and the second largest ventricle assist device program in the country. The LTCF has 201 beds and eight wards including a dementia unit. There was an outbreak of nosocomial cases on one of the wards in the ACH and one of the buildings that contained several wards at the LTCF.

## Methods

To ensure the sampling method could reliably detect environmental RNA before formal sampling began, 100 cm^2^ test surfaces in vacated patient rooms were spotted with 1250 copies of SARS-CoV-2 genomic RNA (BEI Resources). After 10 minutes of drying time, these surfaces were sampled as positive controls. All such controls tested positive using the commercial assay described below.

From 15 May 2020 to 15 Jun 2020, environmental surface sampling and assessments of cleaning thoroughness (adenosine triphosphate (ATP)). and effectiveness (reduction of viral RNA) were performed as previously described [19-20]. Briefly, swabs were streaked over targeted high-touch surfaces for a minimum of 20 seconds in a rolling motion to ensure contact with the entire swab surface. cobas® PCR Media Uni Swabs (Roche Molecular Systems, Inc. Branchburg, NJ) were pre-moistened with the transport media (sterile 40% guanidine hydrochloride Tris-HCl buffer) and analyzed on the cobas® 6800 System (Roche Molecular Systems, Inc. Branchburg, NJ). ATP testing of surfaces was performed using 3M™ Clean-Trace ATP monitoring system.) Sampling at both long-term and acute care facilities was performed using same equipment and in an identical manner by the same authors (DN, LR, EL).

At the ACH and the LTCF, for both random and controlled sampling, dedicated COVID-19 units were used as ‘positive control wards’, and wards which had zero COVID-19 patients identified were uses as ‘negative control wards’. Those that housed both COVID-19 and non-COVID patients were considered ‘mixed wards’. Inpatient dialysis units were considered separately.

### Random Sampling: Assessment of Baseline Environmental Contamination

Eight stationary surfaces sampled in rooms on the outbreak and on the negative and positive ‘control’ wards. These included bed rails, call button/remote control, in-room telephone, over-bed tray table, sink and soap dispenser, chair, windowsill, and the floor. Walking computer workstations, hand-held glucometers, and vital sign machines represented shared surface samples and microwaves and refrigerator samples were taken from the staff common areas. Sampling occurred randomly throughout the daytime hours, without regard to the timing of daily or terminal cleaning. However, since most of these surfaces are cleaned daily, the time from last cleaning to sampling was most often less than 36 hrs.

### Controlled Sampling: Evaluation of Cleaning Effectiveness

At the ACH, immediately after a SARS-CoV-2 infected patient was discharged or transferred out of their room, nursing notified the authors who coordinated with EVS. The same eight stationary surface types were sampled immediately before and after terminal cleaning with a non-bleach sporicidal disinfectant containing hydrogen peroxide and peracetic acid (OxyCide™). Additionally, at the ACH, three types of terminal cleaning enhancements were also evaluated: UV-C treatment at 60,000 mJ/cm2 (RD™ UVC Mobile System), adding electrostatic spraying (Clorox® Total 360®) following terminal cleaning, and adding both UV-C and electrostatic spraying. These enhancements were applied in addition to terminal cleaning when terminal was completed. To ensure each enhancement had an equal amount of RNA contamination present before cleaning, 100 cm2 surfaces in vacated patient rooms were spotted with 1250 copies of SARS-CoV-2 genomic RNA (BEI Resources) before terminal cleaning. During the post-cleaning sample collection, those spotted areas were included in the streaking of the target surface type (floor, bed rail, etc.).

At the LTCF, rooms were sampled before and after routine daily cleaning and also after a professional remediation service using three different proprietary room fogging agents. Adenosine triphosphate (ATP) testing of surfaces was also performed using 3M™ Clean-Trace ATP monitoring system.

The proportion of sampled surfaces with detectible RNA were compared using the 2-sample percent defective tests or Chi squared tests. The mean cycle threshold values of SARS-CoV-2 genes were compared using 2-sample t-test of means.

## Results

### Overall Level of Baseline Contamination

417 surface samples were collected: 214 from the ACH; and 203 from the LTCF. The sampling breakdown and number of positive surfaces is presented in Figures 1 and 2. Of surfaces swabbed, 24% (n = 59) at the ACH, and 23% (n = 47) at the LTCF had detectable RNA. Based on the proportion of positive surfaces, the difference between the overall contamination rates at the ACH and LTCF was not significant (p = 0.313). Similarly, surface contamination based on mean PCR mean cycle threshold values of the *ORF1ab* gene was also similar between the ACH and the LTCF. Mean cycle threshold value at the ACH was 35.4 compared to 35.9 at the LTCF (p = 0.179) (Figure 3).

**Figure 1:**
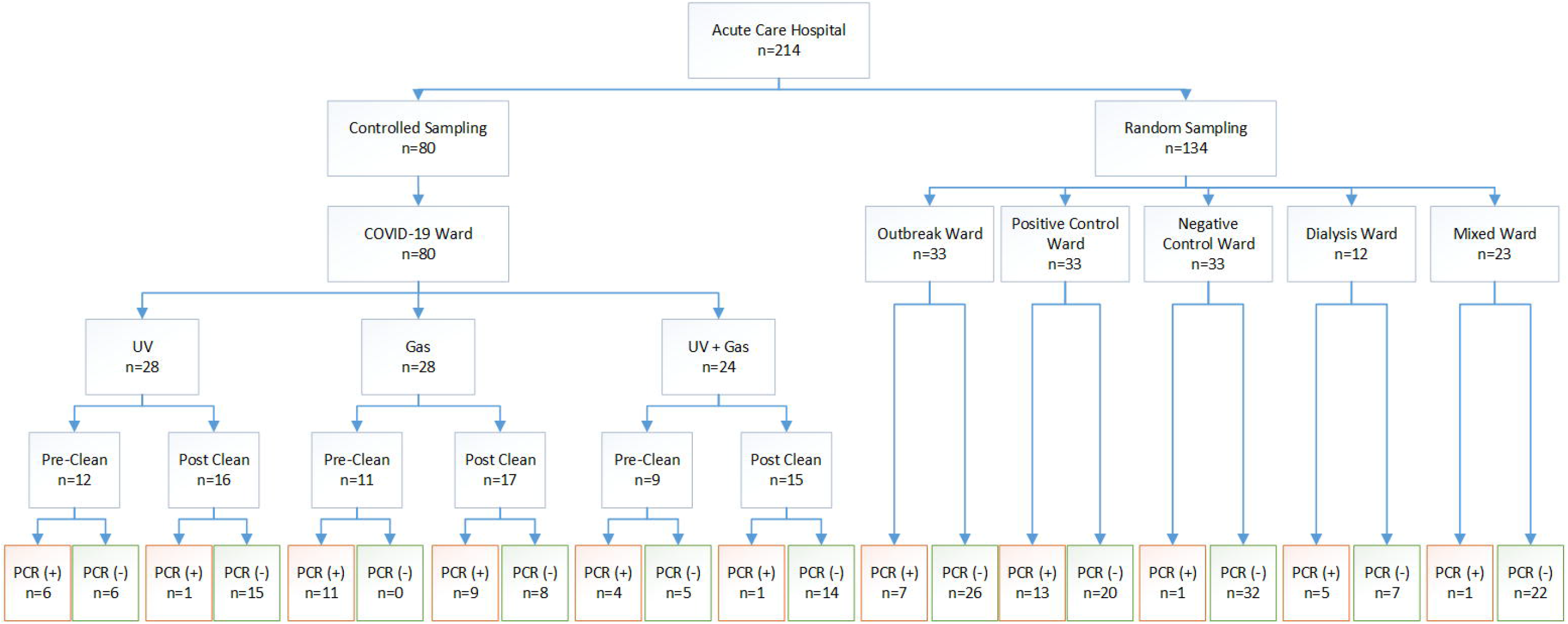
Sample Breakdown at the Acute Care Hospital. Legend: Gas = Electrostatic bleach fogger; UV light = Ultraviolet Light

**Figure 2:**
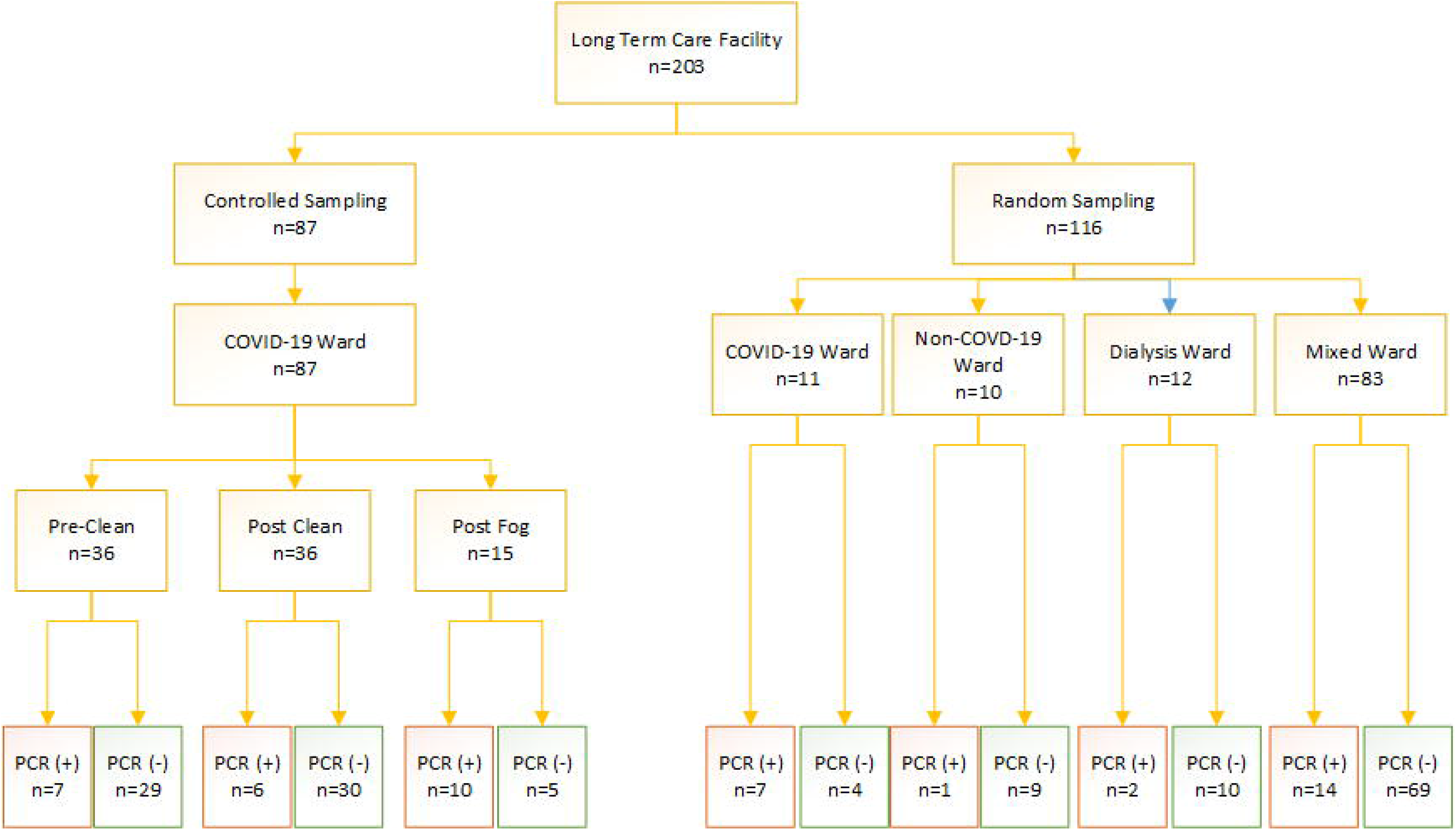
Sample Breakdown at the Long-term Care Facility. Legend: Fog = room spraying with disinfecting chemicals

**Figure 3:**
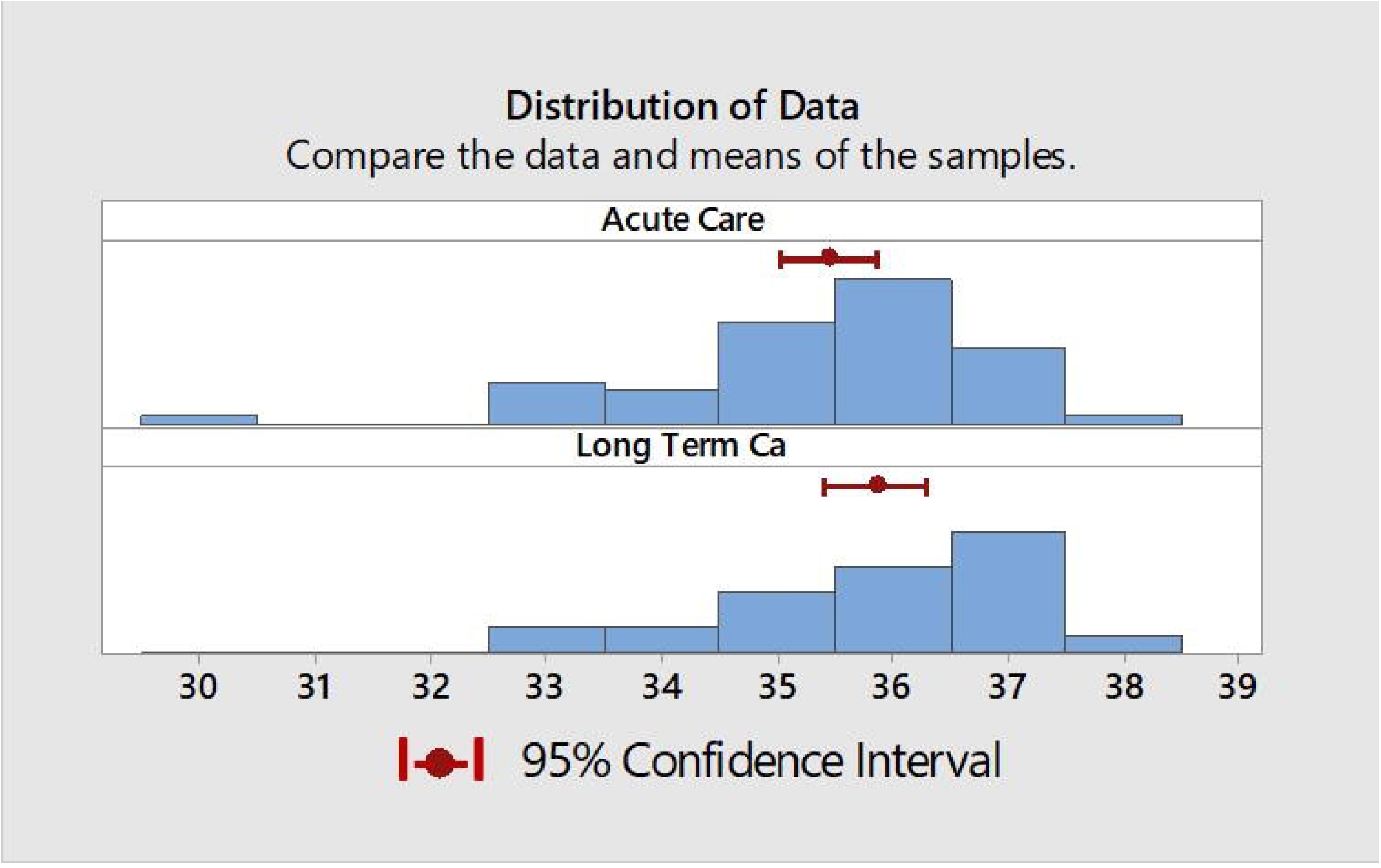
Mean Cycle Threshold Values of ORF1ab at the Acute and Long-term Care Facilities Red = 95% Confidence Intervals

### Randomized Sampling: Contamination on COVID-19, Non-COVID-19, and Inpatient Dialysis Units

At the ACH, 20% of 134 randomly sampled surfaces had detectible RNA (Figure 1). The proportion of surfaces with detectable RNA was not significantly different on the outbreak ward as compared to the dedicated COVID-19 ward (positive control): 7/33 versus 13/33 (p = 0.180). However, there was a significant difference (p = 0.001) when the outbreak ward was compared to the negative control ward, which only had 3% detectable surfaces (1/33).

At the LTCF, 21% of 116 randomly sampled surfaces had viral RNA, which was not significantly different that that seen at the ACH (p = 1.000) (Figure 2). The COVID-19 positive resident rooms had a significantly higher proportion of surfaces with RNA than those of the mixed wards, 7/11 versus 14/83 (p = 0.002). The mixed ward had a higher proportion of surfaces with RNA (17%) than the target of non-COVID-19 ward (10%) (p = 0.035). Stratifying the units further, there was a significant difference between the dementia unit, rehabilitation unit, and the residential unit (p = 0.005) (Figure 4).

**Figure 4:**
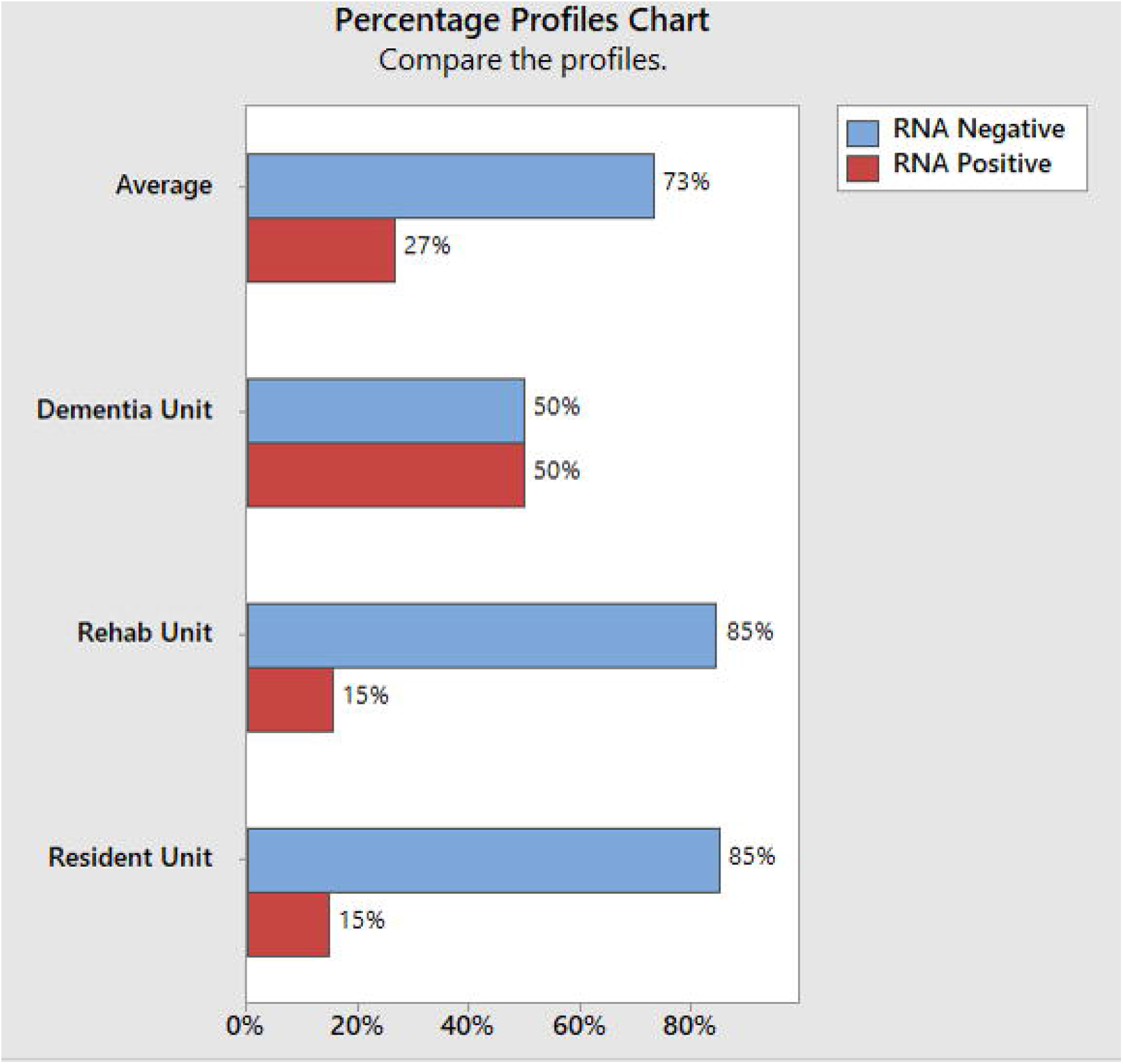
Percentage Profiles of RNA Positive Surfaces at Different Units of the Long-Term Care Facility

Overall (inpatient dialysis at the ACH combined with inpatient dialysis at the LTCF), 29% (7/24) of randomly sampled surfaces had viral RNA. A higher proportion of surfaces were positive at the ACH (5/12) compared to the LTCF (2/12).

### Controlled Sampling: Cleaning Assessments

At the ACH, 437 surfaces throughout the facility including on the outbreak wards, dedicated COVID-19 units, and negative control wards, underwent post cleaning ATP testing as part of routine protocols and quality assessments. The average ATP value was 32 (SD 16.8) RLU. 35 surfaces had both adenosine triphosphate and PCR testing. The mean ATP value of the two PCR positive surfaces was 666 (SD 2.12) RLU, and the mean ATP of 33 negative surfaces was 1274 (SD 2216) RLU; 95% CI for difference -1394 –178) p-value = 0.125.

At the LTCF, 16 surfaces were tested before, and 16 were tested after cleaning. Their mean ATP value was 2308 RLU (SD 2782) (pre-cleaned), and 384 (SD 724; 95% CI 408-3441) post cleaning, p = 0.016.

On the dedicated COVID-19 wards, RNA was detected on 21/32 (66%) of surfaces immediately before cleaning and on 11/48 (23%) of those surfaces immediately after terminal cleaning using a non-bleach sporicidal disinfectant containing hydrogen peroxide and peracetic acid (OxyCide ™) for a 65% post-cleaning reduction (p = 0.001).

UV-C combined with terminal cleaning resulted in an 87% reduction in the proportion of RNA-positive surfaces (50% positive pre-cleaning vs. 6.3% positive after cleaning, p = 0.023) (Figure 1). Terminal cleaning combined with electrostatic spraying (Clorox Pro Total 360 ®) resulted in a 47% reduction (100% vs. 53%, p = 0.010) in detectable target RNA. Spraying plus UV-C led to an 85% reduction (44% precleaned vs. 6.7 % post cleaning, p=0.047) (Figure 5). When UV-C was used, there was zero detectable RNA positive surfaces for the spotted samples (0/12) as compared to electrostatic spraying with 50% of the spotted samples (3/6).

**Figure 5:**
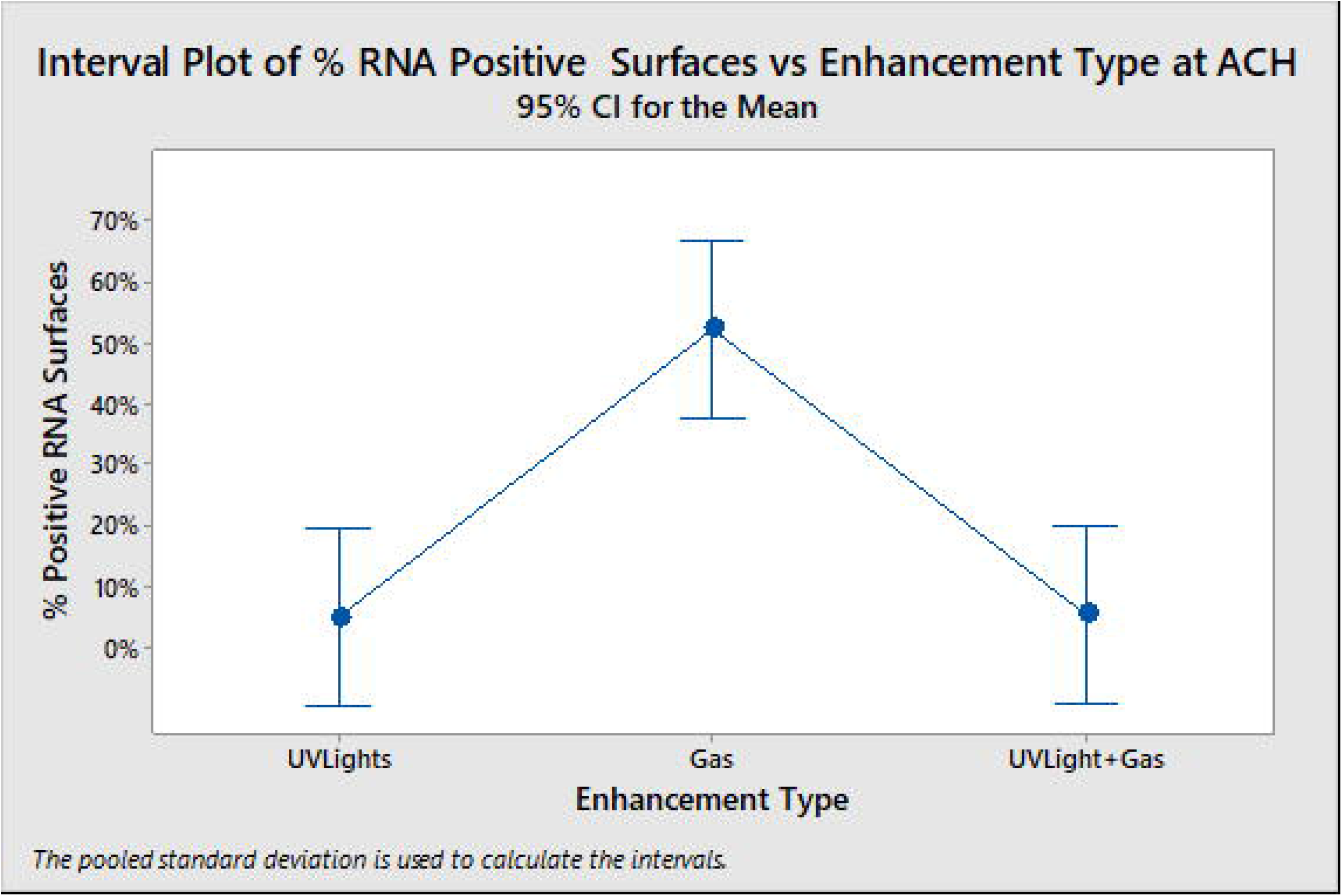
Percent of RNA Positive Surfaces Following Terminal Cleaning Enhanced with Chemical Spraying and Ultraviolet Light. Legend: Gas = Electrostatic bleach fogger; UV light = Ultraviolet Light

At the LTCF, RNA was detected on 10/15 surfaces after professional remediation company fogged patient rooms with a proprietary chlorine dioxide-based disinfectant. In this process, terminal-style surface wiping was not performed. Overall, room fogging combined with terminal-style surface wiping, as routine daily cleaning, led to an 11% reduction in the proportion of positive surfaces. Detectable RNA was present on 19% of surfaces before cleaning versus 17% after cleaning. However, the amount of reduction depended on the time spent wiping and the spraying agent used (Figure 6).

**Figure 6:**
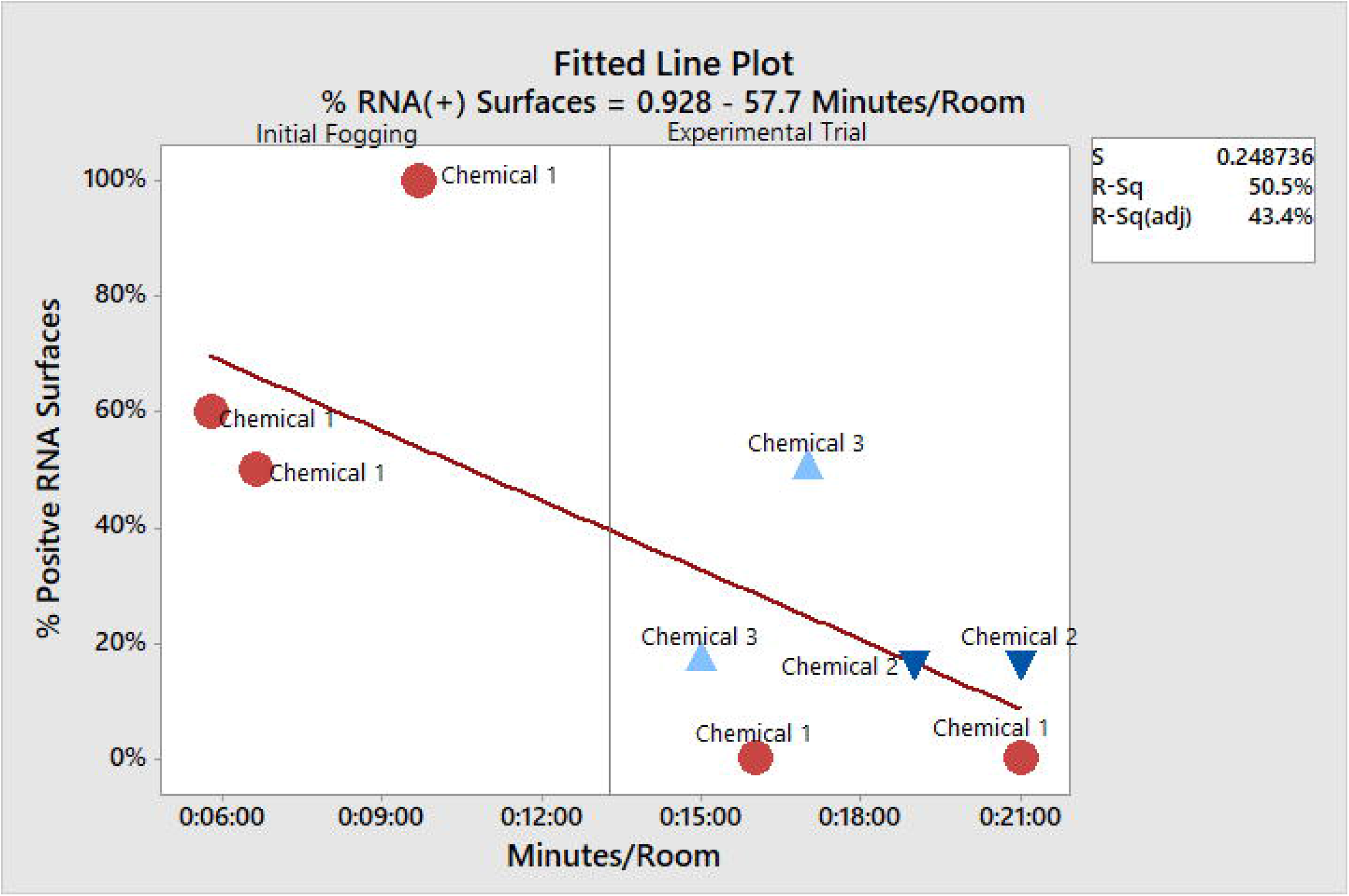
Effect of chemical type and cleaning time on percent of surfaces detected

## Discussion

At both the ACH and LTCF, viral RNA was detected more often on dedicated COVID-19 units, and on wards or units that were experiencing an outbreak or nosocomial transmission. Almost 30% of randomly sampled surfaces in dialysis units, including terminally cleaned dialysis machines, had detectible viral RNA. The overall facility level contamination was no different between an ACH and a corresponding LTCF. The room cleaning protocols used at the ACH significantly reduced environmental RNA, with UV -C light appearing to degrade RNA more than electrostatic spraying. Recognizing: 1) the challenges in cleaning the LTCF because rooms cannot be completely vacated for terminal styled-wiping; and 2) that the ATP values at the LTCF where on average higher than the ACH ATP values, our healthcare system made a considerable investment in UV-C for LTCF.

Commercial cleaning services that spent more time per room cleaning resulted in a larger reduction in contamination, unlike previously published reports that showed time spent cleaning did not translate into more bioburden reduction [21], cleaning by this commercial remediation company that relied on room fogging to be less effective than environmental service employees of the hospital.

Given the scarcity of testing supplies, this study is noteworthy for its sample size. To our knowledge, it is the largest such U.S. study to date. It is also notable for the pragmatic, controlled assessments of cleaning practices under ‘real world’ conditions of staffing shortages during the height of the pandemic. Prior SARS-CoV-2 studies did not have a controlled assessment of cleaning, did not include shared equipment and ATP measurements [3-11].

Our study has several limitations. First, safety concerns and the lack of a BSL-3 level resources precluded the use of viral cultures to determine viability of virus, but nucleic acid of target pathogens can be consequential [19]. Furthermore, lack of culture-based assessments is not unique to our study [4-9]. Second, sponge sticks may be better at capturing pathogens from larger environmental surfaces such as floors and tray tables. These collection devices were unavailable and could not be used in automated PCR reaction tubes. Third, the commercial remediation service and EVS were not compared under identical conditions, but our main objective was not a direct head-to-head comparison of EVS vs contracted remediation service.

Despite these limitations, this report is relevant to nursing homes and hospitals as the findings can assist in allocation of scarce cleaning and disinfection resources for maximum impact. Furthermore, studies of the survivability of the virus on fomites have been criticized as lacking real-life generalizability [22, 23]. Data in this report were obtained from a ‘typical’ general hospital and nursing home, representative of the type of settings (i.e. general/community/non-university-based hospitals) where much of health care is delivered. Studies for these settings may be underrepresented in research because most are carried out in large university-based hospitals.

In summary, surface contamination with SARS-CoV-2 RNA significantly differed based on type of unit, disinfectant, and cleaning regimen. Analysis according to surface, unit, and cleaning regimen, was more informative than facility-level analysis. Additionally, none of the PCR-positive surfaces had Ct values less than 29. Just as patients who remain positive by PCR weeks after infection but with high CTs are not considered to be contagious, the environment viral load we encountered may also place the inanimate surfaces as unlikely source of contagion.

Although the environment may not be a major driver of SARS-CoV-2 transmission at least in community / public settings, the role that environmental contamination play in nosocomial transmission in setting with heavy viral burdens and sick or physiologically vulnerable persons remain less understood. Perhaps another argument for meticulous cleaning of the environment of COVID-19 patients’ rooms is the company that SARS-CoV-2 may keep during the pandemic; e.g., *C. difficle*, MDR-*Acinetobacter*, or MDR-*Candida* that are known to persist in the environment longer than SARS-CoV-2. Further studies are needed to understand how environmental contamination, especially cleaning effectiveness and thoroughness might contribute to SARS-CoV-2 acquisition.

## Data Availability

NA

